# Variability in the spatiotemporal propagation of interictal spikes on magnetoencephalography in temporal lobe epilepsy

**DOI:** 10.1101/2022.12.07.22282951

**Authors:** Daniel J. Zhou, Valentina Gumenyuk, Olga Taraschenko, Bartosz T. Grobelny, Steven M. Stufflebeam, Noam Peled

**Affiliations:** Department of Neurological Sciences, University of Nebraska Medical Center, Omaha, NE; Department of Neurosurgery, Saint Luke’s Health System of Kansas City, Kansas City, MO; MGH/HST Martinos Center for Biomedical Imaging, Charlestown, MA; Harvard Medical School, Cambridge, MA, USA

**Author notes:** **Corresponding Author:** Valentina Gumenyuk, Ph.D., Assistant Professor, Director of the Magnetoencephalography Lab, Department of Neurological Sciences, University of Nebraska Medical Center, 988435 Nebraska Medical Center, Omaha, NE 68198-8435. These authors contributed equally to this work (Daniel J. Zhou, Valentina Gumenyuk, Noam Peled).

**Keywords:** epileptiform discharges, spikes, propagation, magnetoencephalography, temporal lobe epilepsy, temporal plus epilepsy

## Abstract

Magnetoencephalography (MEG) is clinically used to help localize interictal spikes in discrete brain areas through the equivalent current dipole (ECD) method for patients with refractory epilepsy. The propagation of interictal spikes in adjacent regions was shown to be correlated with worse surgical outcomes in patients with temporal lobe epilepsy (TLE). Yet, the ECD method does not account for the temporal dynamics of spike activity, making it challenging to define the margins of the interictal network. Furthermore, the ECD results can be affected by states of arousal or sleep that alter background MEG activity.

In this study, we developed a module in the custom-built Multi-Modal Analysis and Visualization Tool (MMVT) that analyzes the spatiotemporal dynamics of interictal spikes and removes normal variations associated with sleep. We analyzed MEG data from seven TLE patients and characterized their interictal spatiotemporal dynamics. In all patients, the onset of interictal spike activity appeared near the determined site of the epileptogenic zone based on corresponding neuroimaging and EEG data. The propagated source activity by 10 ms ranged from near the region of onset to spreading to extratemporal regions, such as the frontal lobe or insula.

We demonstrated the feasibility of a novel method to assess the spatiotemporal properties of interictal spikes in patients with medically refractory TLE and to better delineate their onset and propagation patterns on MEG analysis. In doing so, we illustrate how variable the spike propagation distribution and speed may be within a 10 ms window. This visualization of the time component of the interictal networks offers an advantage over the currently used ECD method in characterizing the epileptic network and may help better guide surgical planning and provide for improved interventional outcomes in patients with TLE.

## 1. Introduction

Temporal lobe epilepsy (TLE) is often medically refractory and requires invasive therapy resulting in 60-80% seizure freedom after resection or laser ablation of the seizure focus.^1,2^ A reason why some surgeries fail is thought to be due to the epileptogenic activity extending to the extratemporal structures.^3^ The precise localization of the epileptogenic zone (EZ) is critical for improving seizure outcomes, but the process of identifying the margins of EZ remains imperfect.

Interictal epileptiform discharges (i.e., spikes) are strongly associated with epilepsy and contribute to its diagnosis.^4^ Magnetoencephalography (MEG) has demonstrated its value in epilepsy surgery planning by noninvasively detecting interictal spikes and localizing their activity in the deeper cortical regions such as interhemispheric regions and mesial temporal lobe structures.^5–9^ The equivalent current dipole (ECD), which is currently used in clinical practice with MEG, evaluates the burden of spikes in various regions of the brain and locates discrete areas associated with spike activity. However, it does not account for the temporal dynamics of spike propagation, and few studies to date have achieved this using MEG data.^10–12^

The temporal dynamics of interictal activity is highly clinically relevant and correlated with surgical outcomes. In a retrospective study of 37 TLE patients who received surgical resections of the temporal lobe, the patients with temporoparietal spread of propagated interictal spikes on MEG had worse postsurgical outcomes compared to those with spikes restricted to the anterior or mesial temporal lobe.^11^ In another study of 45 patients based on stereo-electroencephalography (sEEG) recordings, the resection of tissue involving source nodes from both the interictal spike onset and propagation was the strongest predictor of good postsurgical outcomes, especially compared to nodes from interictal spike onset alone.^13^

Epileptogenic activity is also known to be potentiated during sleep, and seizure activity is tightly interconnected with sleep function.^14^ The number of interictal spikes quantitatively increases in sleep with a preferential increase in the medial temporal regions.^15^ Normal sleep waveform activity in MEG data, including sleep spindles, vertex waves, and K-complexes, may present as sharp normal transients.^16^ These waveforms could be differentiated from epileptiform spikes by the wave morphology and dipole orientation, with sleep-related activity oriented radially on EEG.^16^ To the best of our knowledge, no previous work has been done to separate the spatiotemporal sources of the interictal activity from the no-spike sleep MEG data.

In the present study, we introduce a new method to remove sleep components from the interictal spatial-temporal dynamics. We then aimed to investigate whether the propagation of the interictal spike activity normalized against sleep activity would better capture the spatial spike distribution in patients with TLE.

## 2. Methods

### 2.1. Subject selection

This is a retrospective study of patients who underwent clinical MEG imaging for the presurgical evaluation of medically refractory TLE at the University of Nebraska Medical Center (Omaha, Nebraska) and Saint Luke’s Clinic (Kansas City, Missouri). The Institutional Review Board of the University of Nebraska Medical Center provided ethical approval, including waiver of informed consent, for the retrospective collection of electronic health record, imaging, and neurophysiologic data for all of the patients in this study (IRB #0714-21-EP). Using the clinical MEG recordings and a novel method developed in our laboratory, we assessed the interictal MEG activity and spatial distribution of spikes in seven patients with TLE. Their clinical and demographic characteristics, as well as findings from their EEG, MRI, and PET imaging studies, were extracted from the electronic medical records.

### 2.2. MEG data recording and processing

The MEG data were recorded in a magnetically shielded room with a whole scalp covered with 306 MEG channels, including 102 magnetometers and 204 planar gradiometers (Elekta Neuromag, Helsinki, Finland). Prior to the MEG study, patients were instructed to reduce habitual total sleep time by ∼40 % by postponing their typical sleep onset by 2-3 hours while keeping their habitual wake time unchanged. During the recordings, patients were positioned supine with head support in the MEG helmet, and all patients achieved stage 2 sleep. The head position was measured and monitored during the entire study via four head indicator coils placed over the EEG cap. The locations of the coils with respect to anatomical landmarks on the head were determined with a 3D digitizer (Polhemus, Vermont, United States). The T1 MRI sequence of the brain was acquired for all patients.

The EEG data from 60 channels were recorded simultaneously using the prefabricated EEG cap (Easy Cap, Germany). The recording bandpass filtering and sampling rate were 0.03-300 and 1000 Hz, respectively. The data were processed offline, and artifacts were removed by spatially filtering the raw data using the temporal extension of Signal Space Separation (tSSS) as implemented in MaxFilter software (Taulu and Simola, 2006). Both MEG and EEG data were recorded for a total of six 10-min sessions, and tracings were reviewed for the presence of epileptiform activity (interictal spikes) by a neurophysiologist (V.G.) and epileptologist (O.T.). Interictal spikes were defined as epileptiform discharge of <70 ms in duration. They were localized using the ECD method provided by the Neuromag software (Neuromag, Helsinki, Finland). The dipoles were accepted when their goodness of fit (into a sphere) was ≥ 80%. Each dipole included the location, orientation, and intensity of current flow in the activated cortical area. All ECDs were superimposed onto the individual patient’s MRI and retained for subsequent analysis of their spatiotemporal spike propagation analysis (Fig. 1-A and 2-A). Ten epochs of waveforms, 4 sec each, were clipped from the raw MEG tracings, 2 sec before and 2 sec after each interictal spike onset. Moreover, ten epochs of sleep stage 2 were clipped for sleep spike-free data, 40 sec in total.

**Figure 1.**
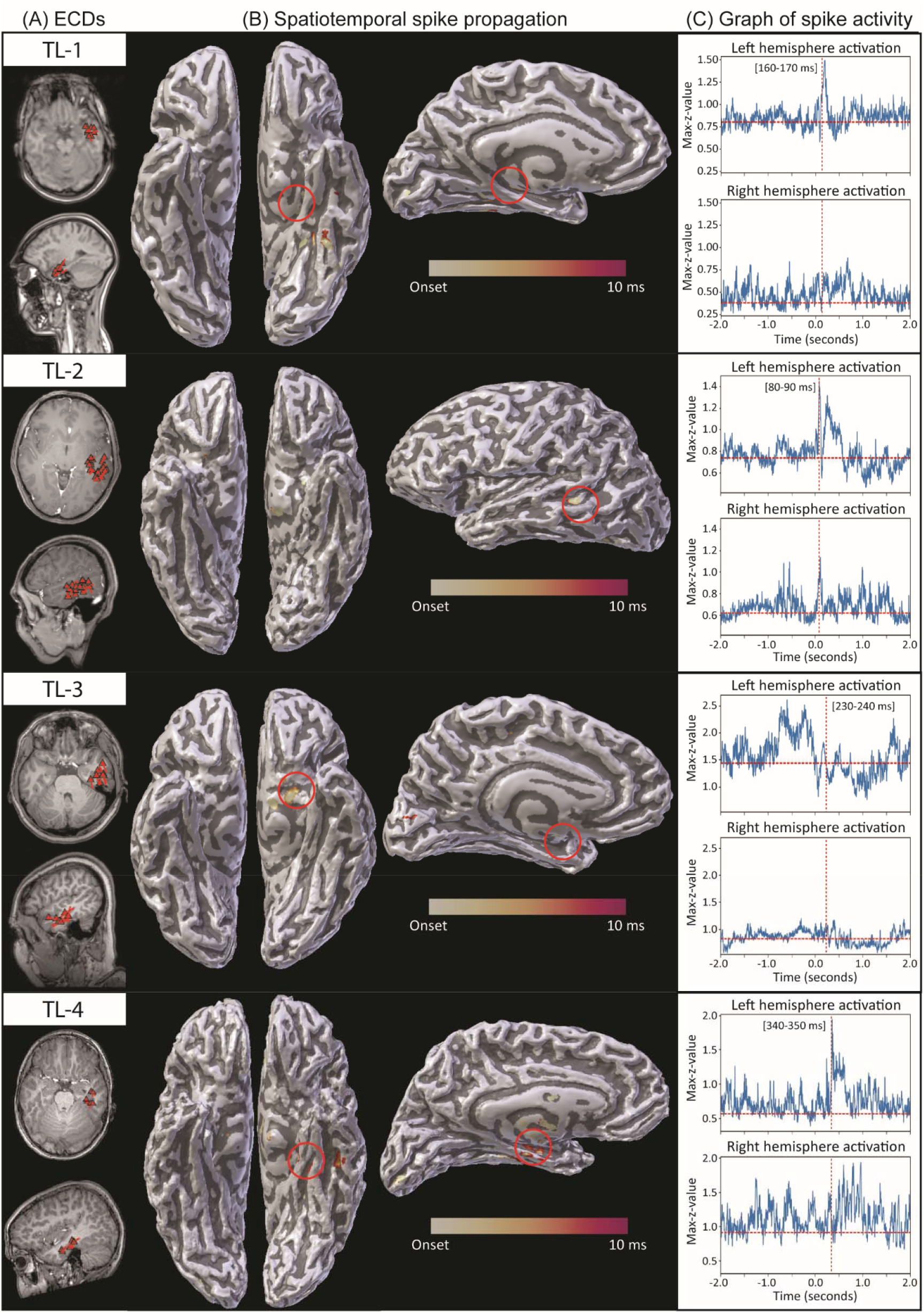
Temporal Lobe (TL) propagation group. Magnetoencephalography (MEG) results in patients with MEG dipole clusters restricted to within the temporal lobe (TL group). Column A shows the MEG clusters using the clinical equivalent current dipole (ECD) method. Column B shows the visualization of interictal spike propagation over time, using the Multi-Modal Visualization Tool (MMVT). The spikes are mapped from 0 (yellow) to 10 ms (red) onto the MRI-derived 3D reconstruction of the patient brain. The region of onset of spike activity is circled in red. Column C shows the average z-value over 4 second epochs, with the onset of spikes selected by the neurophysiologist located at 0.0 seconds. The z-value represents spike burden, based on selected epochs normalized against the patient’s sleep activity without epileptiform discharges. The vertical line represents the selected 10 ms time window based on z-value peak. Each row shows results for each patient, named TL-1 to TL-4.

**Figure 2.**
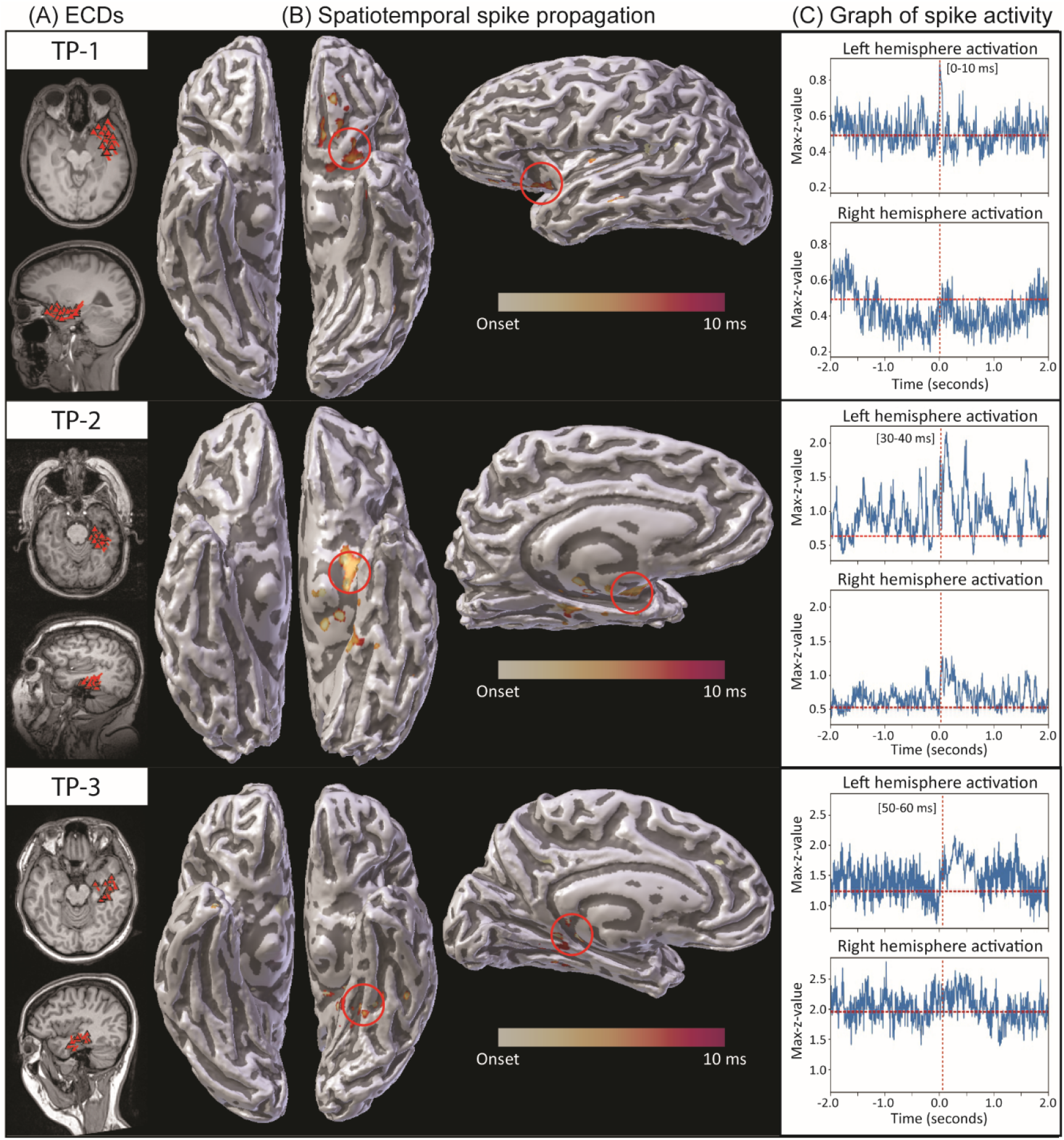
Temporal plus (TP) propagation group. Magnetoencephalography (MEG) results in patients with MEG dipole clusters that are spread beyond the temporal lobe (TP, or temporal plus group). Column A shows the MEG clusters using the clinical equivalent current dipole (ECD) method. Column B shows the visualization of interictal spike propagation over time, using the Multi-Modal Visualization Tool (MMVT). The spikes are mapped from 0 (yellow) to 10 ms (red) onto the MRI-derived 3D reconstruction of the patient brain. The region of onset of spike activity is circled in red. Column C shows the average z-value over 4 second epochs, with the onset of spikes selected by the neurophysiologist located at 0.0 seconds. The z-value represents spike burden, based on selected epochs normalized against the patient’s sleep activity without epileptiform discharges. The vertical line represents the selected 10 ms time window based on z-value peak. Each row shows results for each patient, named TP-1 to TP-3.

### 2.3. Analysis of the interictal spatiotemporal propagation

We first used a Boundary Element Method (BEM) to create a volume conduction model of the head based on individual’s MRI.^17^ Spatiotemporal source distribution was determined using the Dynamic Statistical Parametric Mapping (dSPM) inverse method^18^ from 2 sec before until 2 sec after each ECD. The source distributions were averaged for all ECDs and normalized against stage 2 sleep data containing no epileptiform discharges. The resulting z-values represent the burden of interictal spike activity compared to normal sleep-related activity.

The onset of each interictal spike was defined manually by a neurophysiologist. As a validation for our method, we used the z-values to measure the onset timing objectively and compare that to the onset timings the neurophysiologist found manually. According to our hypothesis, if the z-values measure the burden of interictal activity, we will see a spike in the z-values right after the onset. Moreover, the spike in z-values will occur only in the hemisphere where the dipoles were localized. To test this hypothesis, we first averaged the z-values in space using the Desikan-Killiany-Tourville cortical atlas,^19^ resulting in averaged z-values over time per cortical label. We then selected the maximum value per time point per hemisphere and plotted the results between -2 sec to +2 sec, where 0 is the onset determined manually by the neurophysiologist (Fig. 1-C and 2-C). Using these max-z-values plots, we determined whether we could capture the interictal onsets both in time (onset) and space (lateralization).

In the next step, we examined the spatial-temporal dynamics of the interictal spikes more closely by plotting the z-values on the patients’ cortical surface. As a threshold, we calculated the average value of the max-z-values before the onset for each hemisphere and picked the higher value (red dashed horizontal line in Fig. 1-C and 2-C). We plotted all the z-values above this threshold on the cortical surfaces from the onset to 10 ms using a color gradient from yellow (onset) to red (10 ms). The propagation was plotted layer by layer, starting at 10 ms backward to the onset. A red region denotes spike source data activity (z-values > threshold) only at the end of the selected time window (t = 10 ms). A yellow region denotes the source data activity at or near the onset (t = 0 ms). Using this technique, we could represent the spatial and temporal dynamics of the interictal spikes in one figure (Fig. 1-B, 2-B). As a second validation of our method, we anticipated a region of spike source data activity (z-values > threshold) at the onset where the ECDs were localized.

The model was incorporated into the custom-built Multi-Modal Visualization Tool (MMVT)^20^ to help visualize the spatial and temporal dynamics of spike propagation in the MRI-derived 3D reconstruction of the patient’s brain (Fig. 1-B, 2-B). To validate our method on the spatial domain, we imported the ECDs into the 3D reconstructed brain and compared their locations to the locations where the z-values were above the threshold at the onset.

## 3. Results

We identified seven patients with medically refractory TLE who underwent evaluation with MEG during their presurgical workup (Table 1). In all patients, the ECD results showed dipoles in the temporal lobe areas (Fig. 1-A, 2-A). For all patients, the onset of spikes (i.e., t = 0), defined manually by the neurophysiologist, matched the time of the peak in the max-z-values plots (Fig. 1-C, 2-C, vertical red dashed line). Moreover, the peak was found only in the max-z-values of the hemisphere where the interictal dipoles were localized.

**Table 1.**
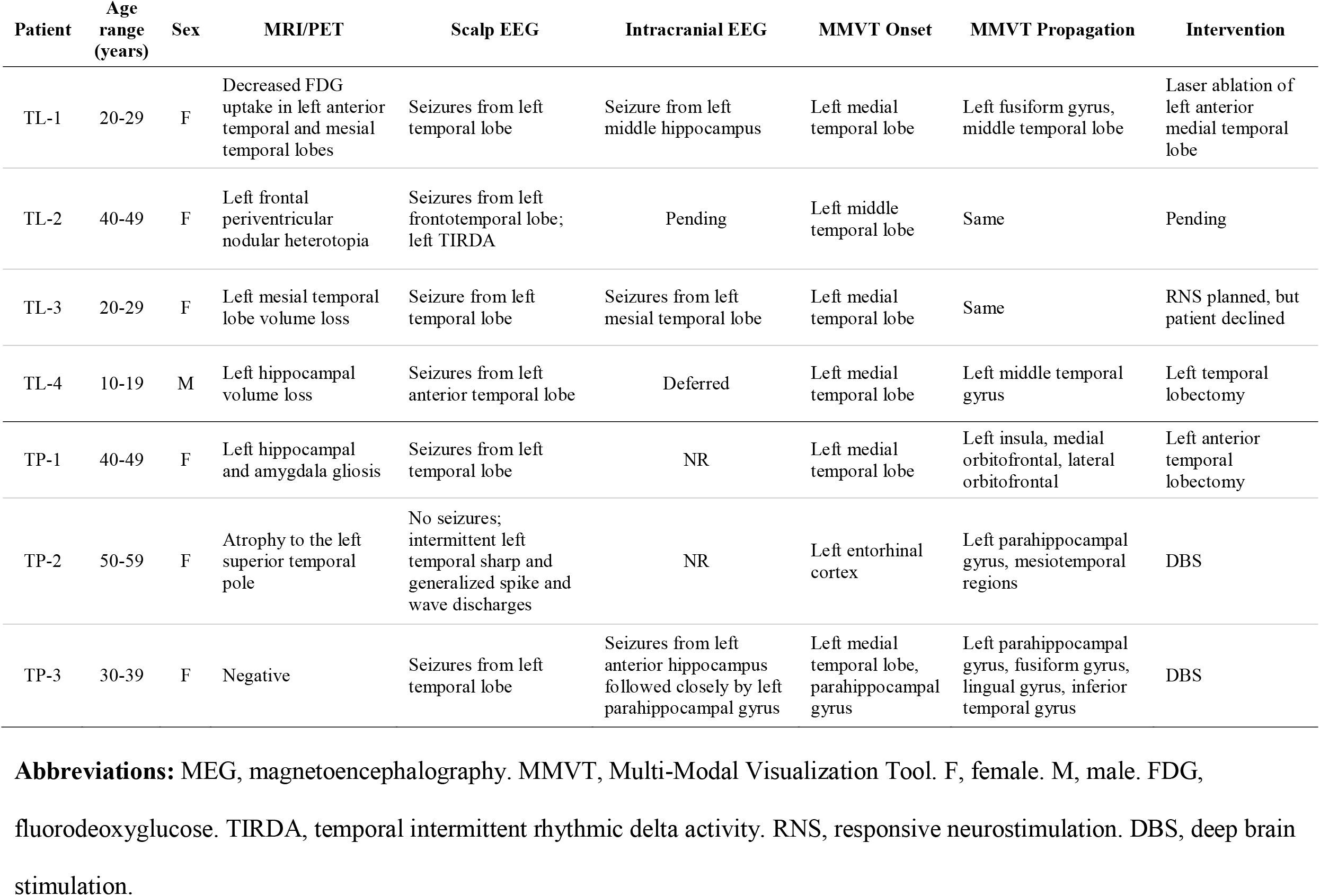
Clinical characteristics of selected patients.

| Patient | Age range (years) | Sex | MRI/PET | Scalp EEG | Intracranial EEG | MMVT Onset | MMVT Propagation | Intervention |
| --- | --- | --- | --- | --- | --- | --- | --- | --- |
| TL-1 | 20-29 | F | Decreased FDG uptake in left anterior temporal and mesial temporal lobes | Seizures from left temporal lobe | Seizure from left middle hippocampus | Left medial temporal lobe | Left fusiform gyrus, middle temporal lobe | Laser ablation of left anterior medial temporal lobe |
| TL-2 | 40-49 | F | Left frontal periventricular nodular heterotopia | Seizures from left frontotemporal lobe; left TIRDA | Pending | Left middle temporal lobe | Same | Pending |
| TL-3 | 20-29 | F | Left mesial temporal lobe volume loss | Seizure from left temporal lobe | Seizures from left mesial temporal lobe | Left medial temporal lobe | Same | RNS planned, but patient declined |
| TL-4 | 10-19 | M | Left hippocampal volume loss | Seizures from left anterior temporal lobe | Deferred | Left medial temporal lobe | Left middle temporal gyrus | Left temporal lobectomy |
| TP-1 | 40-49 | F | Left hippocampal and amygdala gliosis | Seizures from left temporal lobe | NR | Left medial temporal lobe | Left insula, medial orbitofrontal, lateral orbitofrontal | Left anterior temporal lobectomy |
| TP-2 | 50-59 | F | Atrophy to the left superior temporal pole | No seizures; intermittent left temporal sharp and generalized spike and wave discharges | NR | Left entorhinal cortex | Left parahippocampal gyrus, mesiotemporal regions | DBS |
| TP-3 | 30-39 | F | Negative | Seizures from left temporal lobe | Seizures from left anterior hippocampus followed closely by left parahippocampal gyrus | Left medial temporal lobe, parahippocampal gyrus | Left parahippocampal gyrus, fusiform gyrus, lingual gyrus, inferior temporal gyrus | DBS |

**Abbreviations:** MEG, magnetoencephalography. MMVT, Multi-Modal Visualization Tool. F, female. M, male. FDG, fluorodeoxyglucose. TIRDA, temporal intermittent rhythmic delta activity. RNS, responsive neurostimulation. DBS, deep brain stimulation.

Based on their spatiotemporal propagation of source activity over the interval of 10 ms, we separated the patients into two groups: temporal lobe (TL) group, in which the propagation of source spike activity was restricted to the temporal lobe, and temporal plus (TP) group, in which the propagation was distributed over the temporal lobe as well as extratemporal structures (Table 1). The results of the spatiotemporal propagation on the cortical surfaces are illustrated in Fig. 1-B and Fig 2-B for the TL and TP groups, respectively. The propagation is colored using a yellow-to-red gradient, where the onset is in yellow (t = 0) and the propagation areas (closer to 10 ms) are in orange to red. For all patients, there was a spatial cluster on the 3D brain where the ECDs were localized (Fig. 1-A, 1-B circled in red) – a spatial validation for our method.

The spatiotemporal plots (Fig. 1-B, Fig 2-B) were useful in capturing the interictal dynamics at the resolution of 10 ms. Moreover, in reviewing the retrospective clinical data, we found that the z-values spatial cluster fell within the EZ, based on findings from other diagnostic studies performed during presurgical evaluation, including intracranial and scalp EEGs, brain MRI and PET imaging (Table 1).

## 4. Discussion

In the present study, we developed a novel quantitative approach to assess the spatiotemporal dynamics of interictal MEG spikes. Through this tool, we found the onset of interictal spikes in the propagation sequences to be more precisely representative of the EZ in all 7 of the patients. Although MEG was shown to be useful in presurgical planning for epilepsy, the currently employed ECD method lacks the temporal information and cannot detect such dynamics. We demonstrated that MMVT could be applied as a valuable tool for evaluating the spatial and temporal resolution of the interictal activity recorded with MEG. Given the noninvasive nature of the MEG, our method can more precisely localize regions of epileptogenicity and help guide the placement of electrodes for invasive studies, aiding in the determination of appropriate therapeutic interventions.

The surgical treatment for medically refractory TLE is overall effective but fails to provide a seizure-free outcome in 20-40% of the cases.^1–3^ A proposed reason for this is thought to be due to the presence of extratemporal epileptogenic tissue, referred to as temporal plus epilepsy.^21,22^ While this may be possible, there is an alternative thought; a growing recognition that the EZ itself in TLE may be dynamic and evolve over time.^23,24^ Therefore, while the EZ may be characterized at one point through intracranial EEG studies, it may become a different location based on its epileptic network.^23^ Indeed, it was demonstrated that the resection of tissue containing both the interictal spike onset and propagation networks provides a greater chance of seizure freedom compared to targeting the location of spike onset alone.^13^ Similarly, the presence of interictal spike propagation beyond the temporal lobe as demonstrated on MEG was also shown to be associated with worse outcomes.^11^ Still, there are limited data assessing the impact of the propagation velocity of interictal spikes in relation to the accompanying seizure network. We address this by evaluating the source localization of epileptiform activity over a standardized time window of 10 ms, through which we show variability in the distribution of spikes in both locations and amount of spread, despite the ECDs residing in the temporal lobe in both groups.

When localizing the interictal activity recorded during non-rapid eye movement sleep (NREM) state, we wanted to account for the nonepileptic sleep-related brain activity. In the current MEG clinical method, the ECD results can be affected by sleep background when spikes are recorded during sleep.^25^ Therefore, the localization of spike activity would be more accurate when adjusting for normal sleep-related brain activity. We opted to normalize the interictal spike patterns against a sleep background without spike activity, which has not been done before. In this way, we would theoretically focus only on the epileptogenic activity while omitting activity from the patients’ nonepileptic activity.

The limitations of this study include the manual selection of spikes and spike-free sleep epochs by a neurophysiologist and epileptologist. While this approach is similar to the currently used clinical method, it may introduce a selection bias based on the training and experience of the readers. Further, the acquisition technique employed in our laboratory, including sleep deprivation protocol, can deviate from that in other MEG laboratories. Another limitation includes the lack of postsurgical outcome data in these patients, as most of them have completed surgeries within only several months. This limits the definitive conclusions about the clinical utility of this method in predicting seizure outcomes at the present stage.

In summary, we report the feasibility of a novel method to assess interictal spike propagation in patients with drug-resistant TLE. We demonstrated that MMVT could be applied as a valuable tool for evaluating the spatial and temporal resolution of sleep-normalized interictal activity recorded with MEG. Future studies should expand this application to larger cohorts of patients with temporal and extratemporal epilepsy and allow more robust validation of this approach against sEEG data and clinical outcomes. While we do not anticipate that knowledge of interictal spike propagation patterns would allow substitution of the analysis of ictal findings, this method can help provide a better evaluation of the underlying epileptic network and guide intracranial EEG implantation strategies and selection of appropriate interventions.

## Data Availability

All data produced in the present study are available upon reasonable request to the authors

https://github.com/pelednoam/mmvt

https://mmvt.mgh.harvard.edu/

## Acknowledgments

The authors thank Callisto Cordray for her technical assistance in the acquisition of recordings as well as for organizing and navigating the MMVT program files.

## Notes

**Financial Disclosures** N.P. and S.S. are co-founders and shareholders of FIND Surgical Sciences Inc, a startup implementing a clinical decision support platform for epilepsy. O.T. received salary and research support from the NIH P20GM130447 Cognitive Neuroscience and Development of Aging (CONDA) Award. V.G. received research funding from the NIH P20GM130447 Cognitive Neuroscience and Development of Aging (CoNDA) Award.

### Competing Interest Statement

N.P. and S.S. are co-founders and shareholders in FIND Surgical Sciences Inc, a startup implementing a clinical decision support platform for epilepsy

### Funding Statement

O.T. received salary and research support from the NIH P20GM130447 Cognitive Neuroscience and Development of Aging (CONDA) Award. V.G. research funded from NIH P20GM130447 Cognitive Neuroscience and Development of Aging (CoNDA) Award.

### Author Declarations

This is a retrospective study of patients who underwent clinical MEG imaging for the presurgical evaluation of medically refractory TLE at the University of Nebraska Medical Center (Omaha, Nebraska) and Saint Luke's Clinic (Kansas City, Missouri). The Institutional Review Board of the University of Nebraska Medical Center provided ethical approval, including waiver of informed consent, for the retrospective collection of electronic health record, imaging, and neurophysiologic data for all of the patients in this study (IRB #0714-21-EP).

